# Inflammation and cognition in severe mental illness: Patterns of covariation and subgroups

**DOI:** 10.1101/2022.08.25.22279209

**Authors:** Linn Sofie Sæther, Thor Ueland, Beathe Haatveit, Luigi A. Maglanoc, Attila Szabo, Srdjan Djurovic, Pål Aukrust, Daniel Roelfs, Christine Mohn, Monica Bettina Elkjaer Greenwood Ormerod, Trine Vik Lagerberg, Nils Eiel Steen, Ingrid Melle, Ole A. Andreassen, Torill Ueland

## Abstract

Cognitive impairments are common in severe mental illnesses (SMI), such as schizophrenia (SZ) and bipolar (BD) spectrum disorders, with substantial heterogeneity in both diagnostic categories. It has been suggested that dysregulation of immune and inflammatory pathways may contribute to cognitive impairment. This study aimed to investigate covariance patterns between cognitive domains and inflammatory/immune-related markers and further elucidate inter-individual variance in a large SMI and healthy control (HC) cohort (SZ = 343, BD = 298, HC = 770). We applied canonical correlation analysis (CCA) to identify modes of maximum covariation between a comprehensive selection of cognitive measures and inflammatory/immune markers. We found that poor verbal learning and psychomotor processing speed was associated with higher levels of interleukin-18 system cytokines and beta defensin 2, reflecting enhanced activation of innate immunity, a pattern augmented in SMI compared to HC. Applying hierarchical clustering on covariance patterns identified by the CCA revealed a high cognition – low immune dysregulation subgroup with predominantly HC (24% SZ, 45% BD, 74% HC) and a low cognition – high immune dysregulation subgroup predominantly consisting of SMI patients (76% SZ, 55% BD, 26% HC). These subgroups differed in IQ, years of education, age, CRP, BMI (all groups), level of functioning, symptoms and DDD of antipsychotics (SMI cohort). Our findings suggest a potential link between cognitive functioning and innate immune dysregulation.

## Introduction

Schizophrenia (SZ) and bipolar (BD) spectrum disorders are complex severe mental illnesses (SMI) with shared genetic risk factors and neurobiological mechanisms [1]. Cognitive impairments are prevalent and considered a core feature of SMI [2]. These deficits may precede the onset of mental illness [3–5], often persist throughout the illness course [6, 7], and predict poor clinical and functional outcomes [8–11]. Although cognitive impairments are less severe in BD than in SZ, there is substantial heterogeneity within diagnostic categories [12, 13]. It is unclear what underlies the variation in cognitive functioning in SMI, though it is likely due to the complex interplay between genetic susceptibility, biological mechanisms, and environmental factors [14].

One potential biological correlate to cognitive impairment is systemic immune abnormalities such as dysregulated inflammatory pathways. Chronic, low-grade inflammation and immune activation is a risk factor for cognitive impairment in the general population [35–38]. Furthermore, dysregulated inflammatory pathways have been associated with the pathophysiology of SMI [15, 16]. Evidence of immune involvement is supported by genome-wide association studies (GWAS) identifying immune-related genotypes [17–19], and observations of dysregulated levels of inflammatory/immune markers in SMI [16, 20–23]. Such observations have been linked to the more frequent occurrence of somatic comorbidities, particularly cardiovascular disease [24, 25]. Importantly, inflammatory and immune-related processes may influence the central nervous system (CNS) through alteration of the blood-brain barrier (BBB) and modulation of immunocompetent brain cells such as astrocytes and microglia [26–29]. Experimental studies indicate that abnormal glial cell activation may impair neuronal development and homeostasis [30–33]. Thus, it has been suggested that dysregulated immune and inflammatory processes may contribute to cognitive impairments in SMI [34–37].

A recent meta-analysis however, only showed weak associations between immune activation and cognition in case-control SMI studies [38]. Notably, previous studies have focused on individual markers, neglecting the complex interaction between different inflammatory and immune-related signalling systems [39]. Furthermore, recent studies indicate substantial inter-individual differences and potential subgroups of cognitive and inflammatory/immune profiles, which case control studies fail to detect [13, 36, 40]. Such findings are promising as identifying subgroups could help determine who may benefit from personalized treatments.

Recent work has identified subgroups based on comprehensive assessment of inflammatory/immune markers [41–45]. These studies have consistently identified two inflammatory subtypes, with a higher frequency of SMI and healthy controls (HC) in the high and low subtype, respectively. Moreover, the high inflammation subtype in SMI has been associated with poorer response to antipsychotic treatment, greater cortical thickness, and cognitive impairment, but with no differences in symptom severity [36, 41, 43, 44, 46, 47]. Interpretation and clinical relevance is limited due to low sample sizes and the inclusion of few cognitive measures.

Identifying subgroups based on biological data alone could capture variability unrelated to core SMI features such as cognitive functioning. One solution is to investigate immune/inflammatory markers that share variance with cognition [48]. To our knowledge, no previous studies have identified subgroups based on both cognitive functioning and inflammatory/immune markers in SMI. This could help elucidate whether cognitive impairment and elevated levels of immune/inflammatory markers co-occur. This approach has shown merit in SMI studies based on other biological and behavioral data [49, 50], using canonical correlation analysis (CCA), which is a dimension reduction technique that can identify multivariate associations between two sets of variables. Output from CCA can subsequently be used to identify subgroups that have potential clinical relevance [49].

The present study aimed to further elucidate the association between cognitive functioning and inflammation/immune activation in SMI. We performed CCA on a broad range of inflammatory/immune markers and cognitive domains in a large SMI and HC cohort. We applied hierarchical clustering on their patterns of covariance to investigate heterogeneity across diagnostic categories. Here we include nine core cognitive domains that are sensitive to the range of cognitive impairments in SMI [6]. We investigate a large assay of both novel and previously established immune/inflammatory markers associated with SMI. These include markers related to neuroinflammation, BBB integrity, cell adhesion molecules (CAMs), defensins, chemokines, and markers reflecting both adaptive and innate immunity including markers of the interleukin (IL)-18 family as part of the inflammasome system. We address previous concerns related to CCA and clustering techniques [48, 51] by performing cross-validation and stability analyses to evaluate model performance.

## Methods

### Sample

The current study is part of the ongoing Thematically Organized Psychosis (TOP)-study at the Norwegian Center for Mental Disorders Research (NORMENT). Participants meeting the Diagnostic Manual of Mental Disorders (DSM)-IV criteria for SZ or BD spectrum disorders are recruited from psychiatric departments and outpatient clinics in the larger Oslo area. HC from the same catchment area are randomly selected through statistical records and invited by letter to participate. Exclusion criteria for all participants are: (1) outside the age range 18-65, (2) previous moderate to severe head injury, (3) severe somatic or neurological disease interfering with brain functioning, (4) not fluent in a Scandinavian language, and (5) pronounced intellectual deficit (IQ<70). In addition, HC were screened for drug abuse the past 12 months, current or previous history of mental illness, and first-degree relatives with SMI. For the current study, participants with signs of acute infections were excluded (CRP>20 mg/L). The final sample with available cognitive and inflammatory/immune marker data included a total of 1402 individuals with SZ (*n*=343) and BD (*n*=289) spectrum disorders, and HCs (*n*=770). Data was collected between 2004 and 2018. The study was conducted in accordance with the Declaration of Helsinki and approved by the Regional Ethics Committee, and all participants provided written informed consent.

### Clinical assessments

Diagnoses were set by trained clinical psychologists or medical doctors using the Structured Clinical Interview for DSM-IV axis 1 disorders (SCID-I) [52], and included schizophrenia (*n*=175), schizoaffective (*n*=43), schizophreniform (*n*=31), psychosis not otherwise specified (NOS, *n*=94), bipolar I (*n*=173), bipolar II (*n*=103) and bipolar NOS (*n*=14) disorders. Current positive, negative, disorganized, excited and depressive symptom levels were assessed with the Positive and Negative Syndrome Scale (PANSS) [53, 54], and manic symptoms were assessed with the Young Mania Rating Scale (YMRS) [55]. Level of functioning was assessed using the split version of the Global Assessment of Functioning scale (GAF) [56], including symptoms (GAF-S) and function (GAF-F). Age at onset (AAO) of illness was defined as the age of the first SCID-verified psychotic episode for schizophrenia spectrum disorders and manic/hypomanic episode for bipolar spectrum disorders. Duration of illness was estimated by subtracting the AAO from age at assessment. All participants underwent physical examination with blood sampling including measurements of height and weight for calculation of body mass index (BMI). Clinical interviews, physical examination and cognitive testing all occurred within 35 days. The defined daily dose (DDD) of psychopharmacological treatment (antipsychotics, antidepressants, antiepileptics and lithium) was estimated according to guidelines from the World Health Organization Collaborating Center for Drug Statistics Methodology (https://www.whocc.no/atc_ddd_index). Somatic medication use (including anti-inflammatory/immunomodulatory; yes/no) in the SMI group is provided in Supplementary Table 1.

### Cognitive assessments

Cognitive assessment was administered by clinical psychologists (clinical groups) and trained research personnel (HC). We used two test batteries: Battery I (from 2004-2012) and Battery 2 (from 2012-2018). To ensure a comprehensive selection of cognitive domains and the highest possible N, corresponding tests from the two batteries were merged to cover nine domains in addition to intellectual functioning: Intellectual functioning was assessed using the Matrix Reasoning and Vocabulary subtests of the Wechsler Abbreviated Scale of Intelligence (WASI) [57]. *Fine-motor speed* was assessed with the Grooved Pegboard test [58], *Psychomotor processing speed* with the Digit-Symbol Coding task from the Wechsler Adult Intelligence Scale (WAIS-III) [59] or the Digit Symbol task from the MATRICS Consensus Cognitive Battery (MCCB) [60, 61]. *Mental processing speed* (without a motor component) was measured with the color naming and reading subtests from the Color-Word Interference test, Delis Kaplan Executive Functioning System (D-KEFS) [62]. *Attention* was measured using Digit Span forward from the WAIS-III. *Verbal learning* was measured using total recall from the California Verbal Learning Test (CVLT-II) [63], or the Hopkins Verbal Learning Test-Revised (HVLT-R) from the MCCB. *Verbal memory* was measured using long-delay free recall from the CVLT-II, or delayed recall from HVLT-R [64]. For *Semantic fluency* the Category fluency subtest from the Verbal Fluency tests in D-KEFS or MCCB were used. *Working memory* was measured using the total score from the Letter Number Sequencing tests from MCCB or WAIS-III. Finally, *cognitive control* was assessed using the subtests inhibition and inhibition/switching from the Color-Word Interference Test in D-KEFS.

### Peripheral inflammatory and immune markers

Blood was sampled from the antecubital vein in EDTA vials, stored at 4°C overnight, before isolation of plasma that was stored at -80 °C. Average freezer storage time was 6 years (range 1-14), with shorter duration in HC (included as covariate). Markers associated with neuroinflammation included serpin family A member 3 (SA3), alpha-2-macroglobulin (A2M), B-cell activating factor (BAFF), and A proliferation-inducing ligand (APRIL). Neuronal-glial markers reflecting neuroimmune modulation and related to BBB integrity were S100 calcium binding protein B (S100b), furin, glial fibrillary acidic protein (GFAP), neuron specific enolase (NSE/ENO2). The CAMs included were mucosal vascular addressin cell adhesion molecule-1 (MAdCAM-1), junctional adhesion molecule-A (JAMA), intercellular adhesion molecule-1 (ICAM-1), vascular cell adhesion molecule-1 (VCAM-1), and P-selectin (PSEL). The IL-18 system markers analyzed were IL-18 and its binding protein (IL-18BP), as well as IL-18 receptor 1 (IL-18R1), and IL-18 accessory protein (IL-18RAP) reflecting systemic inflammasome activity. The defensins were human neutrophil peptides 1-3 (HNP1-3), beta defensin 1 (BD-1) and beta defensin 2 (BD-2). Lastly, the chemokines included growth-regulated oncogene alpha (GROα/CXCL1), stromal cell-derived factor 1 alpha (SDF1α/CXCL12), eotaxin (CCL11), and regulated upon activation normal T-cell expressed and secreted (RANTES/CCL5). Case-control studies on CAMs, NSE, BAFF, APRIL and IL-18 system components, as well as composite scores based on all markers with overlapping samples have been previously published, [65–69].

Plasma levels of the above biomarkers were measured in duplicate by enzyme immunoassays (EIA) by applying commercially available antibodies (R&D Systems, Minneapolis, MN, USA) in a 384-format using a combination of SELMA (Jena, Germany) and a BioTek (Winooski, VT, USA) dispenser/washer. Absorption was read at 450 nm with wavelength correction set to 540 nm using an ELISA plate reader (Bio-Rad, Hercules, CA, USA). All EIA’s had intra- and inter-assay coefficients <10%. A validation of the stability of the markers regarding effects of diurnal and postprandial variation as well as stability at room temperature and at 4°C have been published previously [69].

### Statistical procedure

#### Data preprocessing

All preprocessing, statistical analyses and visualization of results were conducted in the R-environment (https://www.r-project.org/; v.4.0.3; R-packages reported in Supplementary methods 1). We used a complete-case approach for the cognitive tests which were z-score standardized and some were combined to create the relevant cognitive domain. The inflammatory/immune markers were standardized, outliers removed and replaced with NA using 1.5 x IQR below or above the 25^th^ and 75^th^ percentile, respectively. Missing data on the biomarkers were imputed using Multiple Imputation by Chained Equations (MICE), with predictive mean matching (*m*=5). No variable had >15% missing data (Supplementary Table 2 for missing per variable; Supplementary Figure 1 for MICE output). One-way analyses on plasma levels of the measured inflammatory/immune markers and cognitive domain test scores are found in Supplementary Tables 3-4. Sample and clinical characteristics were compared across groups using permutation (*n*=10000) based t-tests and one-way analysis of variance (ANOVA) for continuous variables, and chi-squared tests for categorical variables.

#### Canonical correlation analysis

We applied canonical correlation analysis (CCA) to identify patterns of covariation between cognitive functioning and inflammatory/immune markers [70]. The new linear combinations (i.e. canonical variates) of the variables generated by the CCA reflect modes of covariance (i.e. canonical variate pairs) between the variable sets. The significance of each mode was assessed by permutation testing (*n*=10000), repeating the CCA on the entire sample for each permutation by randomly shuffling the rows of the inflammatory marker data. The participant loading scores (i.e. mode weights) for the cognitive and inflammatory/immune canonical variates on significant modes were used for interpretation, plotting and in further analyses to investigate the presence of subgroups in the data. Further details on CCA and permutation testing are found in Supplementary methods 2.

#### Out-of-sample cross-validation

To get a more unbiased estimate of the performance of the CCA model in an out-of-sample variable set, we performed a 10-fold cross-validation procedure with 100 repetitions. For each iteration, a new fold was allocated as the test set (20% of the participants), and the remaining 80% of the participants (training set) was submitted to CCA. We then calculated the average canonical correlation from the training set and applied it to the out-of-sample test set to assess generalizability.

#### Stability of canonical loadings

The stability of the canonical loadings (i.e. contribution of each variable on significant modes) was examined following the procedure reported by Dinga and colleagues [48], using their shared R-code at github (https://github.com/dinga92/niclin2019-biotypes). We resampled the data, using their delete-one jack-knife procedure, and replotted the distribution of the canonical loadings for each resample to assess the stability of the loadings.

#### Assessing the influence of covariates on significant modes

Associations between individual loading scores for canonical variates and diagnosis (HC, BD, SZ) were assessed using linear regression, adjusting for age, sex, DDD of psychopharmacological treatments (antipsychotics, antidepressants, antiepileptics and lithium), and BMI, CRP, and freezer storage time (where relevant).

#### Hierarchical clustering

We performed hierarchical clustering to investigate the presence of subgroups in the cognitive and inflammatory/immune canonical variates by generating a distance matrix using the Euclidean distance between the loading scores. To minimize the total within cluster variance, the agglomerative coefficient for several linkage methods (average-linkage, single-linkage, complete-linkage, and Ward’s linkage method) was evaluated. The optimal number of clusters was determined by inspecting the corresponding dendrogram, the elbow method and the average silhouette index. Pairwise-comparisons of clusters with inflammatory/immune marker levels, cognitive domains and demographic characteristics were performed using permutation-based t-tests (Bonferroni corrected).

#### Clustering significance and stability

The significance of the observed silhouette index was tested using a previously reported procedure [48]. Briefly, we first simulated a bivariate Gaussian distribution by taking random samples (*n*=1000) of the covariance matrix for the canonical variates. Next, we applied hierarchical clustering to each random sample and the highest silhouette index was obtained.

We then compared the number of times the silhouette index was smaller for the null distribution, compared to the observations on non-simulated data. Clustering stability was assessed using a bootstrapping resampling procedure. Replicates of the loading scores from the CCA were generated by randomly picking out observations and then replacing them (*n* bootstraps=1000). Hierarchical clustering was performed on each bootstrapped resample. We then computed the Jaccard similarity index ranging from 0-100% and considered an index >0.7 as stable.

#### Code availability

Code will become available upon publication.

## Results

### Sample demographics

Sample demographics are provided in Table 1.

**Table 1.** Sample demographics

|  | **SZ**<br>(N = 343) | **BD**<br>(N = 289) | **HC**<br>(N = 770) | *p*-value | Group comparisons |
| --- | --- | --- | --- | --- | --- |
| Age | 30.2 (10.2) | 32.8 (11.5) | 32.9 (9.1) | $p < 0.001$ | SZ < HC, BD |
| Sex (m/f) | 188/155 | 114/175 | 423/347 | $p < 0.001$ | SZ, HC ~ BD |
| Education (years) | 12.3 (2.3) | 13.6 (2.2) | 14.5 (2.2) | $p < 0.001$ | SZ < BD < HC |
| WASI IQ (2-subtests) | 101 (13.6) | 109 (11.8) | 113 (10.4) | $p < 0.001$ | SZ < BD < HC |
| BMI (kg/m<sup>2</sup>) | 26.3 (5.5) | 25.9 (4.6) | 24.7 (3.7) | $p < 0.001$ | SZ, BD > HC |
| CRP | 3.3 (3.5) | 2.7 (3.0) | 2.2 (2.7) | $p < 0.001$ | SZ > HC, BD |
| PANSS Negative | 13.4 (5.8) | 8.4 (3.4) | | $p < 0.001$ | SZ > BD |
| PANSS Positive | 9.8 (4.2) | 5.8 (2.8) | | $p < 0.001$ | SZ > BD |
| PANSS Disorganized | 5.6 (2.6) | 4.3 (1.6) | | $p < 0.001$ | SZ > BD |
| PANSS Excited | 5.7 (2.1) | 5.2 (1.6) | | $p < 0.001$ | SZ > BD |
| PANSS Depressed | 8.3 (3.3) | 7.7 (2.8) |  | *ns* | - |
| YMRS | 4.3 (4.8) | 3.4 (4.8) |  | *ns* | - |
| GAF Symptom | 45.1 (13.1) | 57.9 (11.2) | | $p < 0.001$ | BD > SZ |
| GAF Function | 45.4 (13.0) | 55.8 (13.1) | | $p < 0.001$ | BD > SZ |
| Age at onset | 23.8 (8.6) | 25.2 (9.5) |  | *ns* | - |
| Duration of illness (years) | 6.7 (8.0) | 7.4 (9.2) |  | *ns* | - |
| Antipsychotics, DDD | 1.0 (0.9) | 0.4 (0.7) | | $p < 0.001$ | SZ > BD |
| Antidepressants, DDD | 0.4 (0.8) | 0.4 (0.8) |  | *ns* | - |
| Antiepileptics, DDD | 0.1 (0.2) | 0.3 (0.5) | | $p < 0.001$ | BD > SZ |
| Lithium, DDD | 0.0 (0.1) | 0.2 (0.5) | | $p < 0.001$ | BD > SZ |
| Total, DDD | 1.5 (1.4) | 1.3 (1.2) |  | *ns* | - |

Presented are means and standard deviations and results from pairwise comparisons.
Abbreviations: HC, healthy controls; SZ, schizophrenia; BD, bipolar disorder; WASI, Wechsler Abbreviated Scale of Intelligence; BMI, body mass index; CRP, C-reactive Protein; PANSS, Positive and Negative Syndrome Scale; YMRS, Young Mania Rating Scale; GAF, Global Assessment of Functioning scale; DDD, defined daily dosage.

### Canonical correlation analysis (CCA)

#### CCA significance and out-of-sample cross-validation

The CCA revealed two significant modes of covariation between the cognitive and inflammatory/immune markers after permutation testing. The first mode had a canonical correlation of .34 (*p* <0.001), and the second mode had a canonical correlation of .22 (*p*<0.001). The null distribution of the canonical correlations from the permutation test is visualized in Supplementary Figure 2. Cross-validation showed that the first mode performed better on unseen data (canonical correlation: mean_training_=0.34, mean_test_=0.26), compared to the second mode where the canonical correlation was substantially lower (mean_training_=0.22, mean_test_=0.09). Due to poor performance of the second mode in the out-of-sample variable set, suggestive of low generalizability, we only considered the first mode moving forward. The variables in each variable set with the largest contributions to the canonical correlation are depicted in Figure 1 A-B.

**Figure 1.**
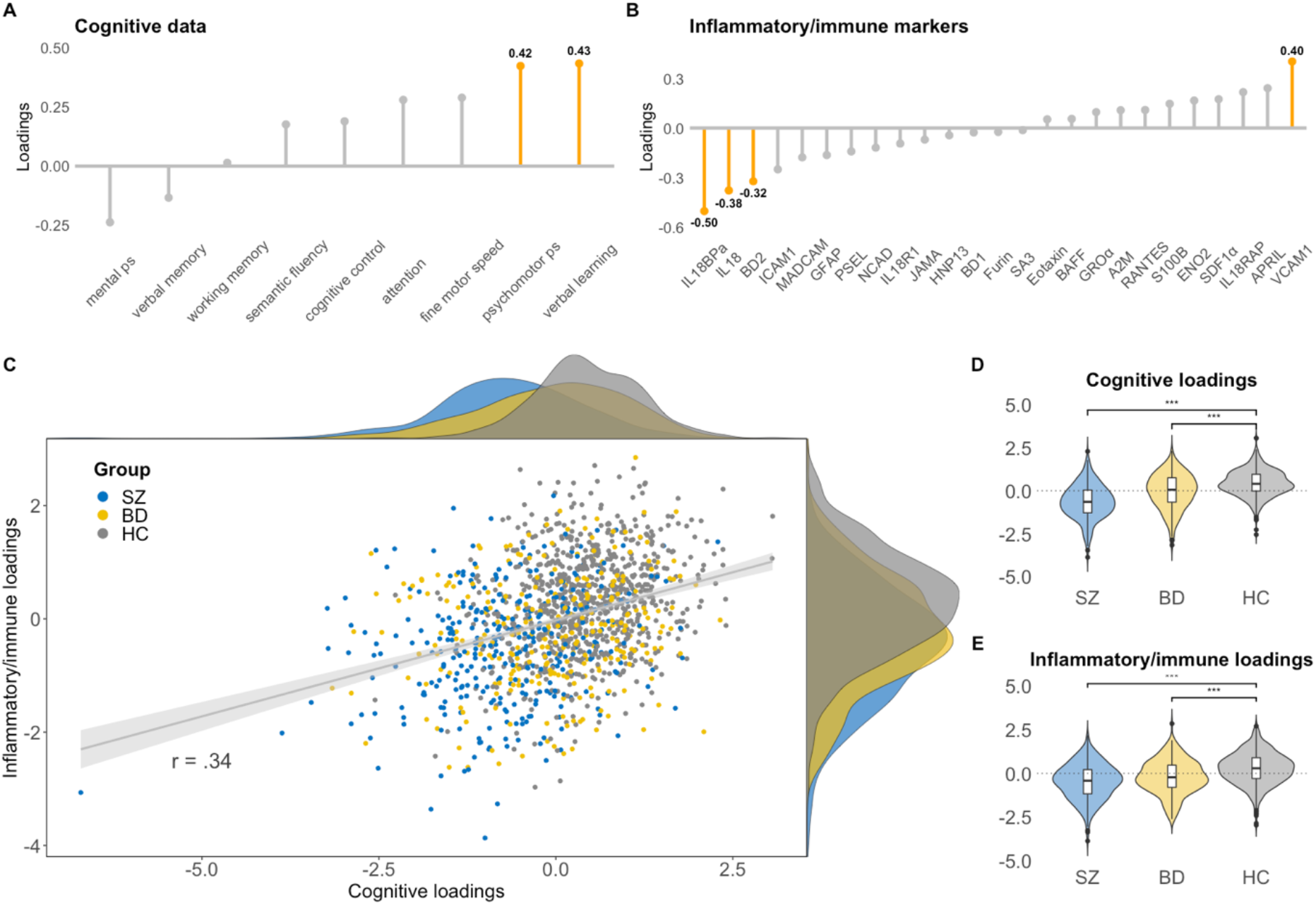
Multivariate mode of covariation between cognitive and inflammatory/immune marker data in SZ, BD and HC. A & B) Shows the highest contributions (in orange) of each variable in the cognitive and inflammatory/immune marker datasets (numbers in bold represents the loading score for the variables with the highest contributions). C) Scatterplot of the individual loading scores, including density plots (top = cognitive loading scores, right = inflammatory/immune marker loading scores). D & E) Violin/box plots showing differences in the loading scores between SZ, BD and HC.

The significant mode of covariation captured verbal learning and psychomotor processing speed which was correlated with a combination of markers of innate immune activation including IL-18, IL-18BP, BD-2 and VCAM-1, with 11% of variance explained. The directionality of the loading scores and the positive correlation indicates that as loading scores decrease on both canonical variates, there is lower cognitive functioning and more severe immune dysregulation (except VCAM-1), including higher inflammasome activation (higher circulating levels of IL-18 system cytokines). Likewise, as loading scores increase, there is higher cognitive functioning and lower degree of immune activation. The scatter plot of the individual loading scores for each canonical variate (Figure 1C), indicates inter-individual heterogeneity and potential subgroups. Small perturbations in the data by leaving one participant out of the CCA did not cause large variations in the canonical loadings, suggesting robust loadings even in the presence of outliers (Supplementary Figure 3).

### Influence of covariates

We next investigated the association between the loading scores for the cognitive canonical variate and diagnosis, while controlling for age, sex and DDD of psychopharmacological treatment (Figure 1C & 1D). Both the BD group (loading score estimate=-0.35±.07, *t*=-4.9, *p*<0.001) and SZ group (loading score estimate=-0.97±.07, *t*=-13.7, *p*<0.001) had significantly lower loading scores on the cognitive canonical variate relative to the HC group. The same pattern was observed for the inflammatory/immune canonical variate for both BD (loading score estimate=-0.41±0.07, *t*=-5.2, *p*<0.001) and SZ (loading score estimate=-0.70±0.08, *t*=-8.5, *p*<0.001), also after controlling for CRP, BMI, and freezer storage time.

### Hierarchical clustering

#### Clustering structure and significance

Out of the linkage methods evaluated, Ward’s came out with the highest agglomerative coefficient (0.99). A 2-cluster solution had the highest average silhouette index (0.37), which was also indicated by visual inspection of the corresponding dendrogram and the elbow method (Supplementary Figure 4). Using a previously described simulation approach [48], the average silhouette index was statistically significant (*p*=0.03), indicating a presence of clusters in the data. The stability analysis suggested a relatively robust cluster assignment, with an average Jaccard similarity Index ∼0.73 (73% overlap) for cluster 1, and ∼0.77 (77% overlap) for cluster 2.

The first cluster identified a subgroup of participants (total *n*=625; SZ=264 [76%], BD=160 [55%], HC=201 [26%]) with negative loading scores on both the cognitive and inflammatory/immune marker canonical variates, whereas the second cluster (total *n*=777; SZ=79 [24%], BD=129 [45%], HC=569 [74%]) was characterized by positive loading scores (all *p*<0.001). The first subgroup (cluster 1) was characterized by higher IL-18, IL-18BP and BD-2 levels, lower VCAM-1 levels, and lower cognitive scores on verbal learning and psychomotor processing speed relative to the subgroup in cluster 2 (all *p*<0.001). See Figure 2 for differences across clusters. Next, we investigated differences between the clusters across demographic and clinical data (SMI only). The subgroup in cluster 1 had lower IQ and years of education, and higher age, CRP, and BMI, relative to the subgroup in cluster 2 (all *p*<0.001). Among the participants with SMI, the subgroup in cluster 1 had lower functioning (GAF-S, GAF-F), more positive, negative, and disorganized symptoms and used a higher DDD of antipsychotics compared to the subgroup in cluster 2 (all *p*<0.001). See Table 2 for comparisons.

**Table 2.** Differences between subgroups on demographic and clinical data

|  | **Cluster 1**<br>**(N = 625)** | **Cluster 2**<br>**(N = 777)** | ***p* value<sup>A</sup>** | **95% CI<sup>B</sup>** | **Cluster comparisons** |
| --- | --- | --- | --- | --- | --- |
| Age | 33.2 (11.1) | 31.4 (8.9) | $p < 0.001$ | [0.8, 2.9] | Cluster 1 > Cluster 2 |
| Sex (m/f) | 370/255 | 355/422 | $p < 0.001$ | - | Cluster 1 ~ Cluster 2 |
| Education (years) | 12.9 (2.3) | 14.4 (2.3) | $p < 0.001$ | [-1.7, -1.2] | Cluster 1 < Cluster 2 |
| WASI IQ (2 subtests) | 103.7 (13.1) | 114.1 (9.9) | $p < 0.001$ | [-11.7, -9.1] | Cluster 1 < Cluster 2 |
| BMI (kg/m<sup>2</sup>) | 26.5 (5.0) | 24.4 (3.7) | $p < 0.001$ | [1.5, 2.5] | Cluster 1 > Cluster 2 |
| CRP | 3.1 (3.3) | 2.2 (2.7) | $p < 0.001$ | [0.6, 1.2] | Cluster 1 > Cluster 2 |
| **SMI group** | **BD = 160**<br>**SZ = 264** | **BD = 129**<br>**SZ = 79** |  |  |  |
| PANSS Negative | 11.9 (5.6) | 9.7 (4.6) | $p < 0.001$ | [1.3, 3.1] | Cluster 1 > Cluster 2 |
| PANSS Positive | 8.6 (4.4) | 6.8 (3.3) | $p < 0.001$ | [1.1, 2.4] | Cluster 1 > Cluster 2 |
| PANSS Disorganized | 5.3 (2.4) | 4.3 (1.7) | $p < 0.001$ | [0.6, 1.4] | Cluster 1 > Cluster 2 |
| PANSS Excited | 5.5 (2.0) | 5.3 (1.8) | ns | [-0.2, 0.5] | - |
| PANSS Depressed | 8.1 (3.1) | 7.9 (2.9) | ns | [-0.3, 0.7] | - |
| YMRS | 4.0 (4.9) | 3.6 (4.4) | ns | [-0.4, 1.2] | - |
| GAF Symptom | 48.8 (13.6) | 55.6 (13.3) | $p < 0.001$ | [-0.9, -4.5] | Cluster 1 < Cluster 2 |
| GAF Function | 47.73 (12.9) | 55.2 (14.9) | $p < 0.001$ | [-9.8, -5.1] | Cluster 1 < Cluster 2 |
| Age at onset | 24.9 (9.6) | 23.4 (7.8) | ns | [-0.0, 3.0] | - |
| Duration of illness (years) | 7.5 (9.2) | 6.0 (7.0) | ns | [-0.0, 2.9] | - |
| Antipsychotics, DDD | 0.8 (0.9) | 0.5 (0.8) | $p < 0.001$ | [0.1, 0.4] | Cluster 1 > Cluster 2 |
| Antidepressants, DDD | 0.4 (0.9) | 0.4 (0.7) | ns | [-0.1, 0.2] | - |
| Antiepileptics, DDD | 0.2 (0.4) | 0.2 (0.4) | ns | [-0.1, 0.1] | - |
| Lithium, DDD | 0.1 (0.3) | 0.1 (0.4) | ns | [-0.1, 0.0] | - |
| Total, DDD | 1.5 (1.3) | 1.2 (1.3) | ns | [0.1, 0.5] | - |

Presented are means, standard deviations and results from permutation t-tests (and chi-square for sex) comparing clusters on demographic and clinical data. *Note:* HC, healthy controls; SZ, schizophrenia; BD, bipolar disorder; WASI, Wechsler Abbreviated Scale of Intelligence; BMI, body mass index; CRP, C-reactive Protein; PANSS, Positive and Negative Syndrome Scale; YMRS, Young Mania Rating Scale; GAF, Global Assessment of Functioning scale; DDD, defined daily dosage. <sup>A</sup>Bonferroni corrected *p* values (sample characteristics 0.05/5; clinical characteristics 0.05/15). <sup>B</sup>95 percent (Monte-Carlo) permutation percentile confidence interval of the mean difference.

**Figure 2.**
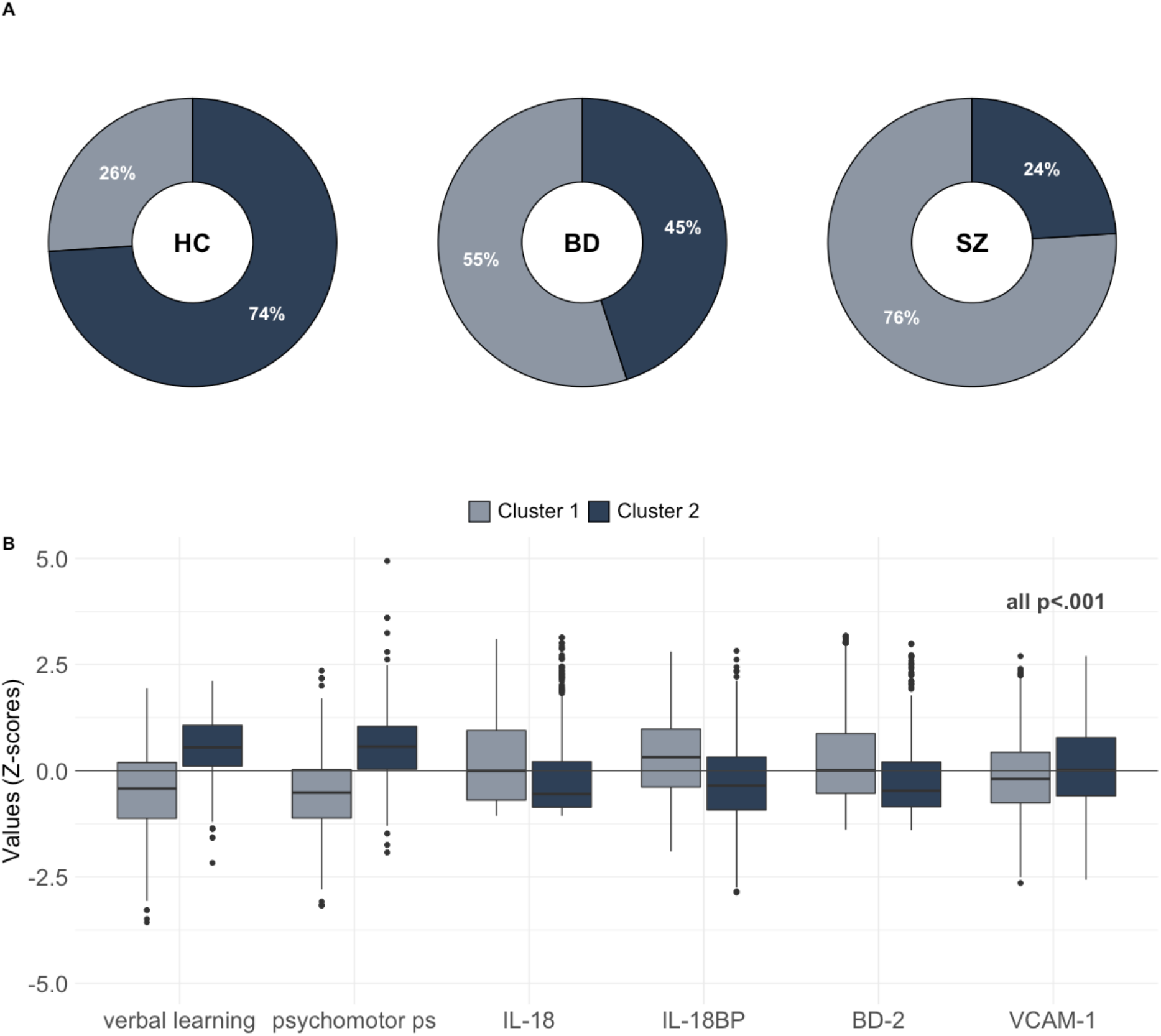
Hierarchical clustering of CCA data in SZ, BD and HC. A) Percentage of SZ, BD and HC in cluster 1 and cluster 2. B) Boxplots showing differences between the subgroup cluster 1 and cluster 2 on the cognitive domains and inflammatory/immune markers identified from the CCA.

## Discussion

In a large SMI and HC cohort, we identified shared covariance between verbal learning and psychomotor processing speed and markers of innate immune activation, including IL-18, IL-18BP, BD-2, and VCAM-1. Furthermore, the covariance patterns indicated two subgroups with distinct cognition – immune dysregulation with differing demographics and clinical severity. Our findings suggest innate immune activation and cognitive impairment co-occur in a subgroup predominantly consisting of SMI, highlighting the importance of considering inter-individual variance in future research.

The cognitive domains that shared covariance with markers of innate immune activation, verbal learning and psychomotor processing speed, are among the most affected cognitive domains in SZ and BD [71–76]. Impairments are evident in clinical high-risk individuals with subsequent conversion to SMI [77] and potentially qualify as endophenotypes in both SZ and BD [71, 78]. A recent meta-analysis demonstrated modest correlations between IL-6 and it’s downstream mediator CRP and impairment in both of these domains [38]. However, CRP is a non-specific marker of systemic inflammation. Our findings, present also in adjusted analysis controlling for BMI and CRP, suggest that more specific markers reflecting other pathogenic processes, such as activation of innate immune responses, may be relevant for cognitive functioning.

Based on our findings we can begin to speculate about potential inflammatory/immune-related mechanisms that may influence cognitive functioning. IL-18 system components regulate innate immune responses and are broadly expressed by neurons, astrocytes and microglia, and may influence permeability of the BBB and induce neuroinflammatory states [79]. We have recently demonstrated increased levels of these IL-18 system components in SMI, associated with increased gene expression of the inflammasome components NLRP3 and NLRC4 in circulating immune cells [67]. The inflammasome is a key innate immune system function that is associated with many human diseases [80]. A growing number of studies suggest that inflammasome activation can influence cognitive functioning, particularly in autoimmune and neurodegenerative diseases [80–83]. In addition, experimental studies have shown promise in mitigating cognitive impairment by inhibiting inflammasome activation, which could be a potential treatment target for several pathologies [84].

Similar to IL-18, the small antimicrobial peptide BD-2, mainly produced by neutrophils and epithelial cells as well as macrophage cells, plays an important role in regulating innate immune responses. While representing a protective component against bacterial, viral and fungal infections, defensins may cause collateral damage in host cells by disrupting cellular membranes, and have been shown to diffuse across the BBB [85]. Dysregulated expression of BD-2 in microglia and astrocytes has been suggested to prolong dendritic cell activity, which could mediate release of pro-inflammatory cytokines ultimately promoting loss of neuronal function and impacting cognition [86]. Based on the increased BD-2 levels indicated by the CCA, we speculate that similar mechanisms could be relevant in SMI. BD-2 has pleiotropic effects, acting as a chemokine binding to CCR6 with effects on T cells and dendritic cells [87], linking innate (inflammation) and adaptive (lymphocyte activation) immune responses. Furthermore, BD-2 induces IL-18 release in keratinocytes [88] and conversely, IL-18 may trigger BD-2 release in innate cells such as macrophages [89].

While we recently reported similar levels of sVCAM-1 in SMI and HC [90], our finding that low sVCAM-1 was associated with cognitive impairment could indicate an alternative role for the soluble form of this protein. VCAM-1 may mediate adhesion of monocytes, lymphocytes, and neutrophils to the vascular endothelium including immune cell trafficking via the BBB [91]. While increased VCAM-1 expression is a key marker for endothelial activation during vascular/systemic inflammation [92], inflammatory challenge may enhance shedding of VCAM-1 from human brain endothelial cells [93]. Additionally, *in vitro*, sVCAM-1 may act as a competitive inhibitor of ligand binding, blocking leukocyte adhesion to activated human brain endothelial cells [94]. Chronically elevated levels of circulating IL-18 may also downregulate sVCAM-1 in both immune and non-immune cells [95]. Taken together, we speculate that chronic dysregulation of innate immune regulatory loops in SMI could enhance systemic IL-18 signaling, together with BD-2 and sVCAM1 expression, impacting BBB permeability and neuroinflammation, thereby influencing cognition.

We identified two groups reflecting heterogeneity in cognitive functioning and inflammatory/immune status in SMI and HC. While there was a larger proportion of SMI participants in the more compromised group (low cognition – high immune dysregulation subgroup), they were also represented in the less compromised subgroup. We additionally found a proportion of HC in the compromised group sharing several characteristics with SMI. This is in line with findings of inter-individual variance in cognitive functioning and inflammatory/immune status in SMI and in the general population [96, 97].

Importantly, our approach suggests that SMI individuals with lower cognitive functioning and higher immune/inflammatory dysregulation may experience more symptoms, worse functioning and have higher DDD of antipsychotics. Symptom severity has not previously been linked with subtypes based on inflammatory markers alone [36, 41, 43, 44, 47]. Additionally, we found no differences between subgroups for AAO or DOI, which could suggest a common cognitive and inflammatory/immune activation pattern independent of illness stage. This supports findings of cognitive deficits and increased inflammation in both first-episode psychosis and chronic illness [41, 99]. Notably, differences across studies based on the selection bias of inflammatory/immune markers and cognitive outcomes could contribute to differing observations from subgroup-studies.

Some limitations should be acknowledged. While cognitive functioning and immune/inflammatory markers can be influenced by multiple factors, an open question is whether they simply co-occur or whether there is a causal relationship between them. Due to the cross-sectional design and measures of peripheral inflammatory/immune markers we are unable to draw conclusions regarding causality. Longitudinal studies, and evaluation of the inflammatory and immune markers in CNS (i.e. cerebrospinal fluid) are needed to clarify this. The study has several strengths including a robust methodology, a transdiagnostic approach, a large sample, and a comprehensive selection of cognitive domains and a large inflammatory/immune screening assay. Cross-validation, stability analyses, and evaluation of the cluster solution, further strengthen our findings, although replication in independent datasets is needed.

In conclusion, we identified patterns of covariance between cognitive functioning and inflammatory/immune markers, linking poor verbal learning and psychomotor processing skills to increased innate immune activation markers, including IL-18 system cytokines and BD-2, with the strongest associations in SMI. Based on covariance patterns we identified two subgroups of cognitive functioning and inflammation associated with differing patterns of functioning and symptom levels that transcended diagnostic categories. Our findings suggest that the IL-18 system, and perhaps inflammasome activation, could be an interesting path for future investigation of cognitive impairment in SMI.

## Data Availability

The data are not publicly available due to privacy and ethical restrictions.

## Acknowledgements

This study was funded by the South-Eastern Norway Regional Health Authority (grant number 2020089) and Research Council of Norway (#223273).

