## Supplementary figures for "Inflammation and cognition in severe mental illness: Patterns of covariation and subgroups"

### 1. MICE outputs

**A**

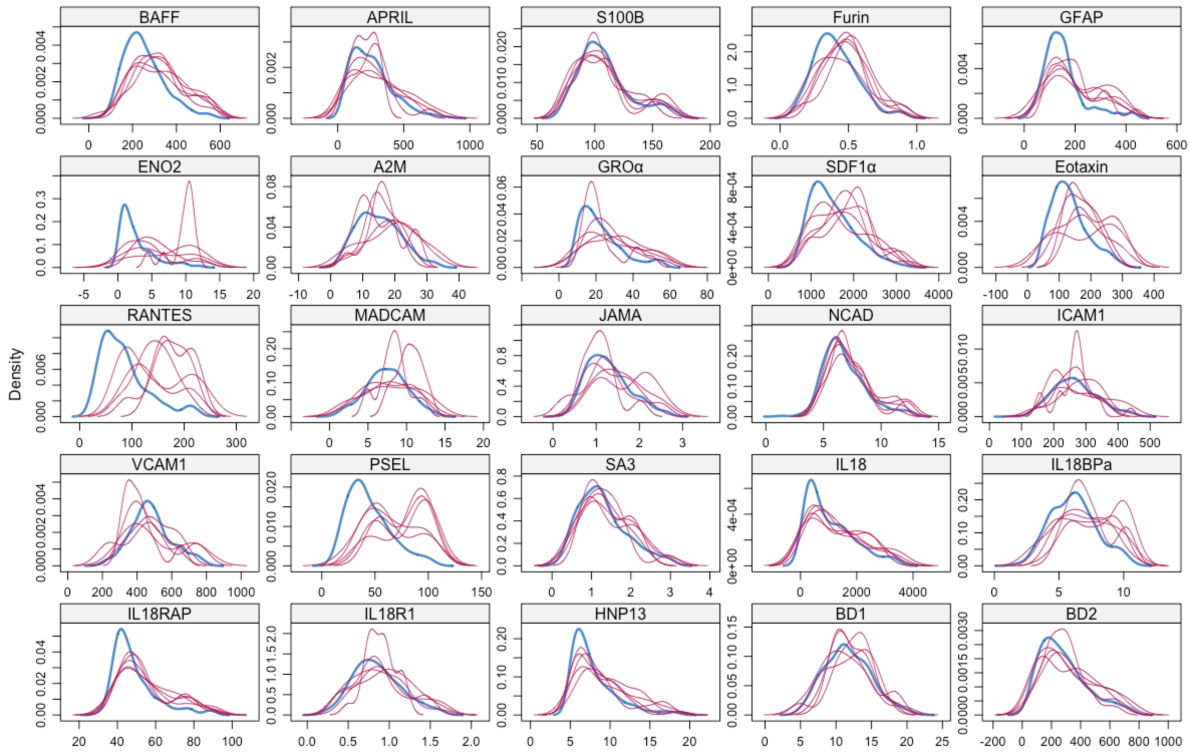


**B**

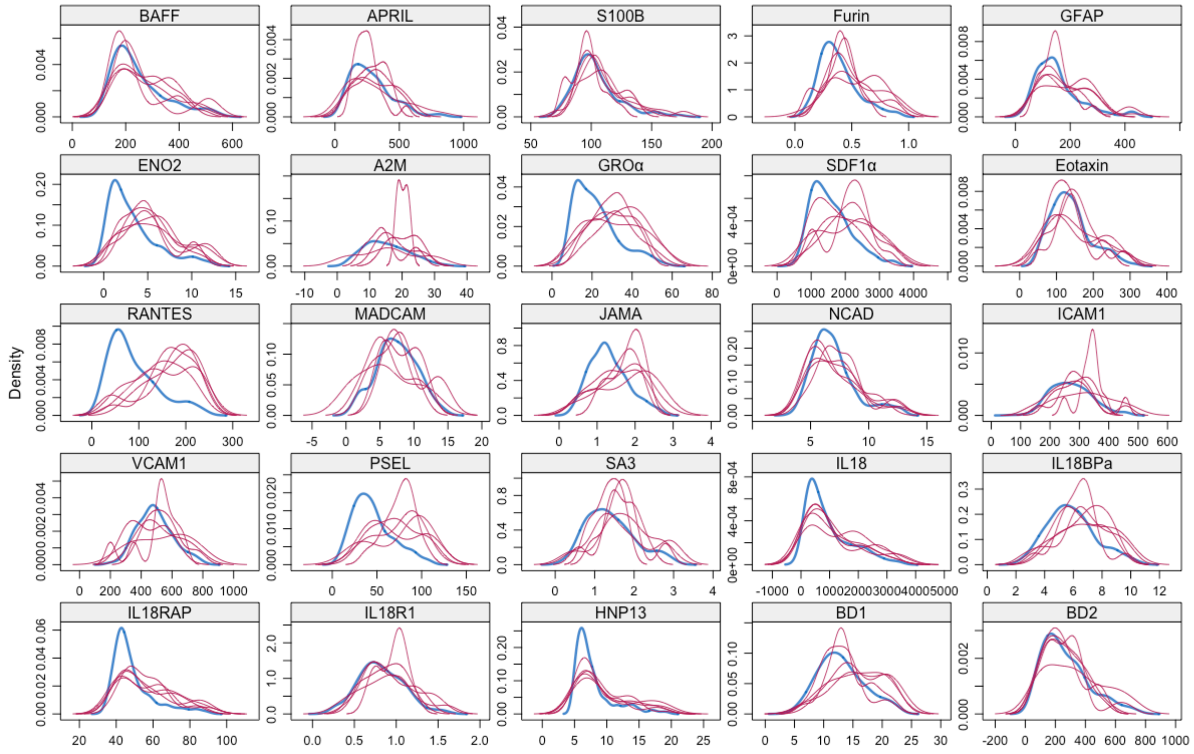

**C**

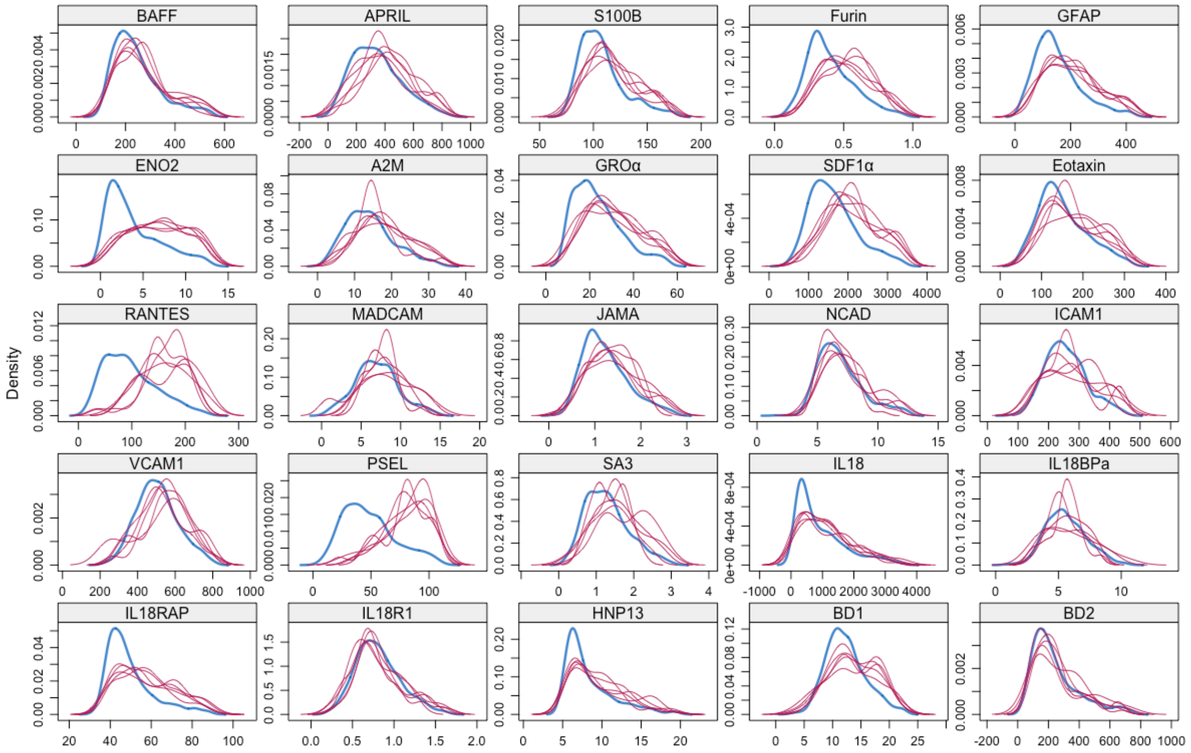


**Supplementary Fig. 1 (A-C)** Shows the density of the imputed data for each imputed dataset (in magenta) and the observed data (blue). Imputations (*m*=5) with similar distributions as observed data is the best fit, we selected imputation dataset 1. MICE imputation was completed separately for A) the SZ group, B) the BD group, and C) the HC group.

### 2. Results from permutation test of CCA

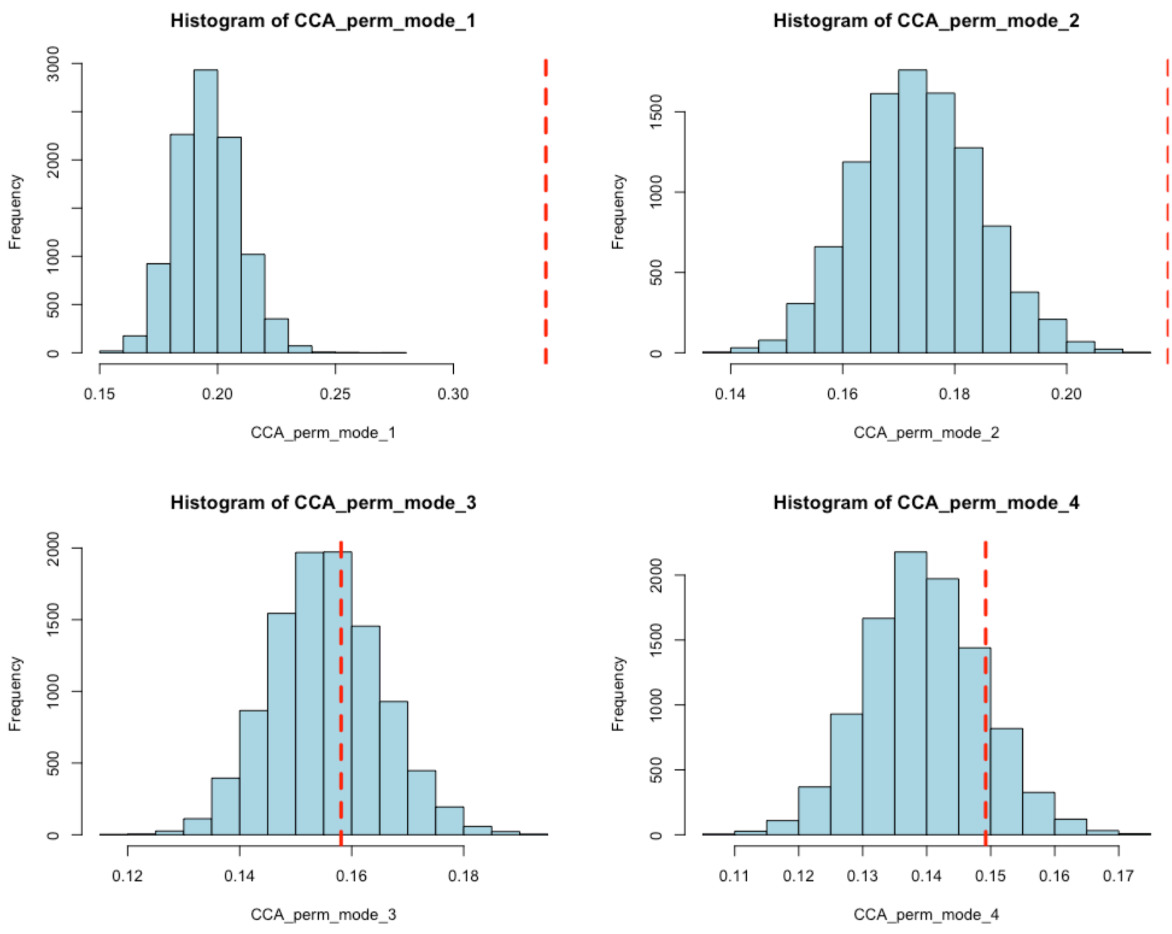


**Supplementary Fig. 2** Shows the null distribution from the permutation test of the CCA canonical correlation for the first 4 out of 9 possible modes. The red dotted line is the empirical canonical correlation. The first and the second mode was significant at $p$<0.001 (top panels).

### 3. Stability of canonical loading scores

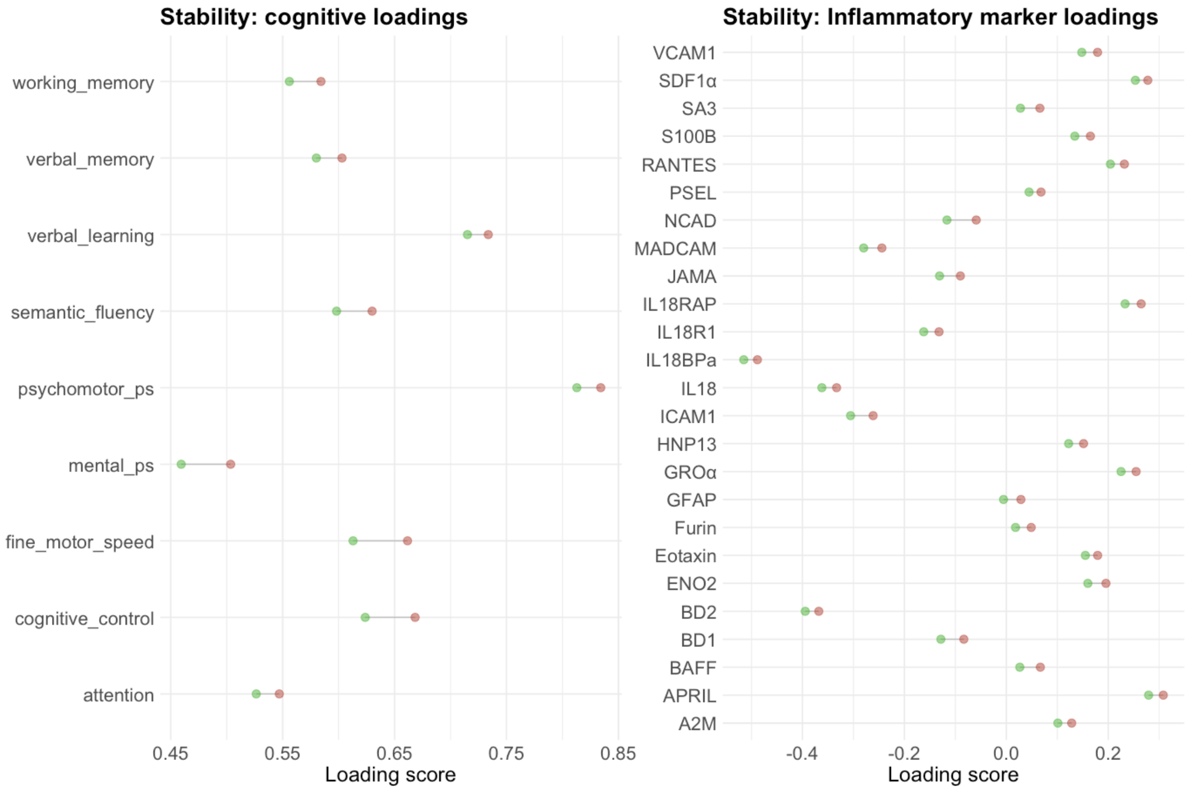


**Supplementary Fig. 3** Shows results of the stability analysis based on Dinga et al., (2019) available R code at github (https://github.com/dinga92/niclin2019-biotypes). Both the cognitive and inflammatory/immune marker loadings were stable when running the CCA and leaving one participant out of the analysis.

### 4. Results of hierarchical clustering analysis

A

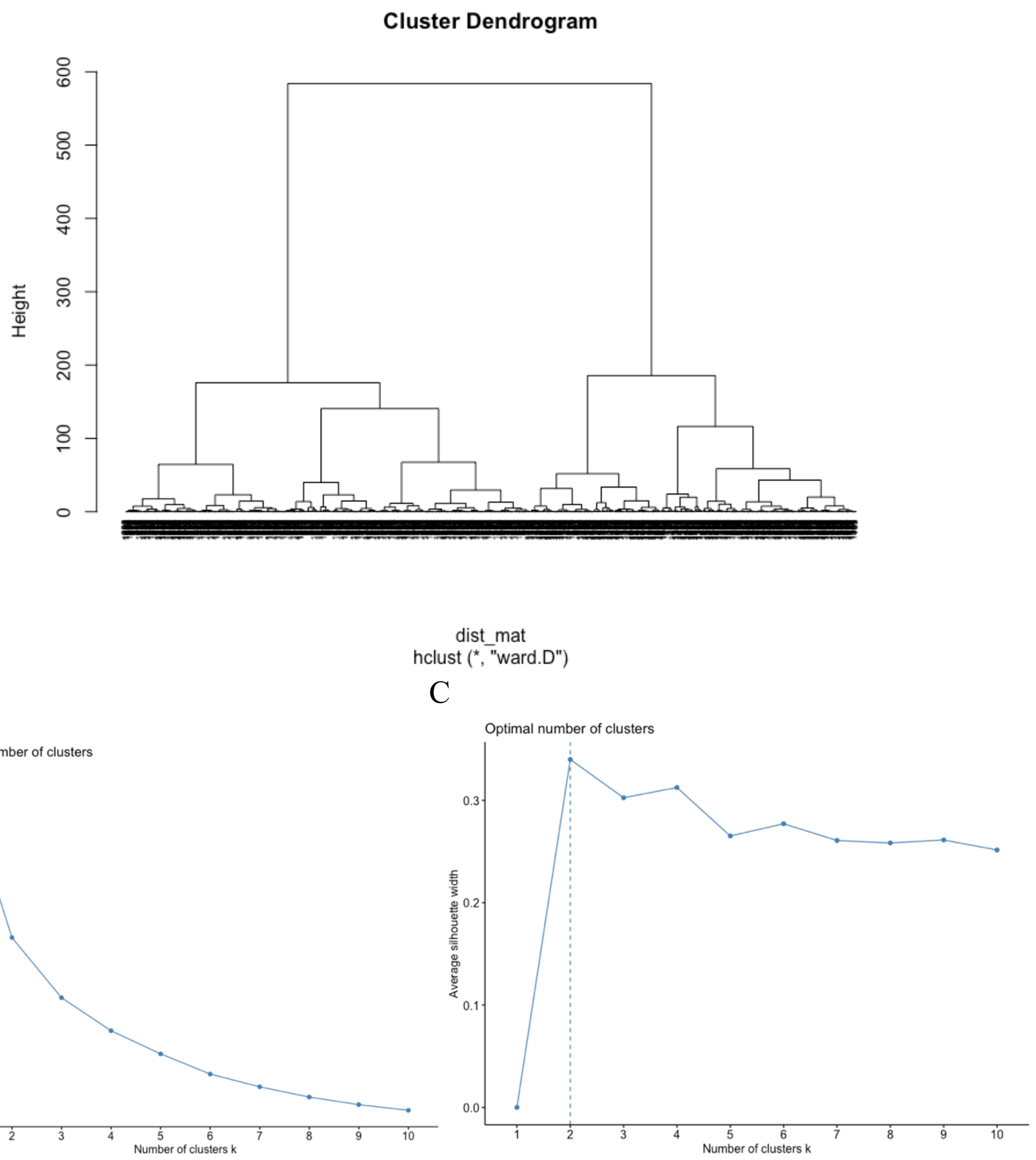


**Supplementary Fig. 4 (A-C)** Evaluation of optimal number of n clusters for hierarchical clustering. A) Dendrogram based on “hclust”. B) Elbow method, C) Average silhouette index.
