## Supplementary methods for "Inflammation and cognition in severe mental illness: Patterns of covariation and subgroups"

### 1. R-packages used for visualization and main analyses

- For visualization the R-package “ggplot2” was applied [1].
- Multiple Imputation by Chained Equations (MICE) was applied using the R-package “mice” [2].
- The R-packages “MKinfer” [3] and “rcompanion” [4] were used for sample and clinical characteristics (permutation based t-tests and ANOVAs).
- The CCA and permutation test was implemented using R-packages “candisc” [5] and “rsample” [6].
- The 10-fold cross-validation procedure was implemented using the R-package “caret” [7].
- Clustering was performed using R-packages “cluster” [8], “dendextend” [9] and “factoextra” [10].
- Clustering stability was assessed using the R-package “fpc” [11].

### 2. CCA, permutation testing and stability analysis

CCA is related to dimension reduction techniques, like principal component analysis (PCA). CCA seeks to find linear combinations of variables from two datasets that are maximally correlated with each other [12].These new linear combinations, called canonical variates, form modes (similar to components in PCA; canonical variate pairs) of covariation, and the correlation between them is called a canonical correlation. The maximum number of modes that can be extracted from the data equals the number of variables in the smallest dataset (in our case there were nine possible modes). Several modes of covariation can be significant if the subsequent mode is uncorrelated with previous modes. To assess the significance of each mode, we performed permutation testing. The CCA was repeated for each permutation by randomly shuffling the rows of the inflammatory markers matrix such that they no longer corresponded to rows in the cognitive data matrix, thereby breaking the association between the two datasets. We performed 10000 permutations, which generated a null distribution of canonical correlations. This procedure effectively controls for multiple comparisons across all possible modes. However, the significance of CCA modes is traditionally assessed by

Wilks Lambda test statistics. When assessing this statistic, we only found one significant mode, although permutation testing identified two (the second mode had low generalizability by cross-validation and was not followed-up further).

The subject-variable-ratio (SVR) in CCA is important. A recent methodological study found that for large samples (n ~1000) it is recommended to perform dimension reduction if SVR is very low, i.e. n variables is very large (>50)[13]. When *n* variables are considerably below the sample size (i.e. high SVR) the CCA correlation strength and stability is found to be robust. Due to the high SVR in our study, we did not perform dimension reduction prior to CCA. However, we did assess the stability of our CCA findings with stability analysis of the canonical loadings under slight changes of participants (leave-one-out procedure). The distribution of the canonical loadings for each resample was then plotted to assess the stability of the loadings. If there is a large variation in the canonical loadings under small perturbations of the data, then the CCA model is unstable and unlikely to improve under more extensive resampling procedures [14]. In addition, the resampling can help identify whether outliers have significant impact on the canonical loadings.
