## Supplementary tables for "Inflammation and cognition in severe mental illness: Patterns of covariation and subgroups"

**Supplementary table 1.** Somatic medication use by SMI group

| **Somatic medications** | **SZ (N = 343)** | **BD (N = 289)** |
|---|---|---|
| Anti-inflammatory/immunomodulatory, N (%) | 11 (3.2) | 1 (0.3) |
| Antidiabetics, N (%) | 5 (1.5) | 1 (0.3) |
| Cardiovascular/lipid modifying, N (%) | 15 (4.4) | 10 (3.5) |
| Antihistamines, N (%) | 17 (4.9) | 9 (3.1) |
| Gastrointestinal drugs, N (%) | 13 (3.8) | 7 (2.4) |
| *Other, N (%) | 51 (14.9) | 64 (22.1) |

* Other includes vitamins, minerals, analgetics, thyroid agents, pulmonary agents, urological agents, musculoskeletal agents, contraceptives, sex hormones, anxiolytics, anti-inflammatory (local administrative agents, i.e. crème/inhalators), hematological agents, parenteral nutrition agents, substance dependency agents and mucolytic agents.
Abbreviations: Schizophrenia (SZ), Bipolar disorder (BD), Healthy controls (HC)

**Supplementary table 2.** Percentage missing per inflammatory/immune marker

| Inflammatory/immune markers | SZ (N = 343) | BD (N = 289) | HC (N = 770) | Total (N = 1402) |
|---|---|---|---|---|
| | | Percentage (%) missing | | |
| BAFF | 10.2 | 10.3 | 9.6 | 9.9 |
| APRIL | 4.6 | 6.9 | 5.4 | 5.5 |
| S100B | 12.2 | 8.3 | 9.8 | 10.1 |
| Furin | 7.8 | 5.1 | 6.6 | 6.6 |
| GFAP | 12.2 | 10.7 | 12.3 | 11.9 |
| ENO2 | 3.2 | 4.4 | 7.5 | 5.8 |
| A2M | 4 | 1.3 | 3.1 | 2.9 |
| GROα | 4.6 | 5.5 | 5.8 | 5.4 |
| SDF1α | 7.2 | 5.5 | 8.8 | 7.7 |
| Eotaxin | 2.6 | 4.1 | 4.6 | 4 |
| RANTES | 2.6 | 4.8 | 3.7 | 3.7 |
| MADCAM | 4.3 | 2.4 | 2 | 2.7 |
| JAMA | 3.2 | 3.4 | 5.9 | 4.7 |
| NCAD | 12.8 | 9.3 | 8.5 | 9.7 |
| ICAM-1 | 6.9 | 3.4 | 1.6 | 3.3 |
| VCAM-1 | 2.6 | 3.1 | 2.9 | 2.9 |
| PSEL | 2.9 | 4.1 | 3.3 | 3.4 |
| SA3 | 4.6 | 3.1 | 3.5 | 3.7 |
| IL-18 | 9.3 | 10 | 7 | 8.2 |
| IL-18BP | 9.6 | 5.5 | 2.5 | 4.9 |
| IL-18RAP | 12.2 | 11.7 | 13.7 | 12.9 |
| IL-18R1 | 5.8 | 4.8 | 3.8 | 4.5 |
| HNP13 | 11 | 12.1 | 14.1 | 12.9 |
| BD-1 | 4.6 | 6.5 | 5.7 | 5.6 |
| BD-2 | 11.3 | 8.9 | 6.1 | 7.9 |

*Note:* Missing data was imputed using Multiple Imputation by Chained Equations (MICE). See Supplementary figure 1 (A-C) for density distributions of observed and imputed data.
Abbreviations: Schizophrenia (SZ), Bipolar disorder (BD), Healthy controls (HC)

**Supplementary Table 3.** Inflammatory/immune-related marker levels between SZ, BD and HC

| **Immune/Inflammatory markers**[a] | **SZ (N = 343)** | **BD (N = 289)** | **HC (N = 770)** | ***p*-value**[b] | **Effect size**[c] | **Pairwise comparisons**[b] |
|---|---|---|---|---|---|---|
| *Neuroinflammation* | | | | | | |
| A2M (μg/mL) | 14.6 (9.3) | 14.3 (10.0) | 13.5 (8.6) | **0.04** | 0.003 | ns |
| APRIL (pg/mL) | 235.5 (196) | 249.7 (204) | 326 (237) | **<0.001** | 0.06 | BD, SZ<HC |
| BAFF (pg/mL) | 238 (123) | 215 (119) | 226 (112) | **0.04** | 0.003 | ns |
| SA3 (μg/mL) | 1.2 (0.8) | 1.3 (0.8) | 1.2 (0.8) | **0.01** | 0.005 | HC, SZ<BD |
| *BBB integrity* | | | | | | |
| ENO2 (ng/mL) | 1.8 (3.1) | 2.6 (3.4) | 3 (4.7) | **<0.001** | 0.03 | SZ<BD, HC; BD<HC |
| Furin (ng/mL) | 0.4 (0.2) | 0.3 (0.2) | 0.4 (0.2) | **0.03** | 0.003 | BD<SZ, HC |
| GFAP (pg/mL) | 139 (78.5) | 139 (100) | 139 (107) | ns | - | - |
| S100B (ng/mL) | 104 (26.1) | 101.2 (21.8) | 104 (26.1) | **<0.01** | 0.006 | BD<SZ, HC |
| *Chemokines* | | | | | | |
| Eotaxin (pg/mL) | 122.4 (74.9) | 131 (66.1) | 138.5 (81.6) | **<0.001** | 0.02 | SZ, BD<HC |
| GROα (pg/mL) | 19.5 (13) | 20.4 (14.7) | 21.7 (14.9) | **<0.01** | 0.006 | SZ<HC |
| RANTES (ng/mL) | 77 (56.3) | 74.2 (73.3) | 90.3 (76) | **<0.001** | 0.01 | SZ, BD<HC |
| SDF1α (pg/mL) | 1437 (716) | 1497 (816) | 1577 (799) | **<0.01** | 0.008 | SZ<HC |
| *Cell adhesion molecules* | | | | | | |
| ICAM-1 (ng/mL) | 260.3 (95) | 268.6 (99) | 249.7 (94) | **<0.01** | 0.006 | HC<BD |
| JAMA (ng/mL) | 1.2 (0.7) | 1.2 (0.7) | 1.1 (0.7) | **0.02** | 0.004 | HC<BD,SZ |
| MadCAM-1 (ng/mL) | 7.5 (3.4) | 7.3 (4.1) | 6.8 (3.6) | **<0.01** | 0.007 | HC<SZ |
| NCAD (ng/mL) | 6.4 (2.4) | 6.7 (2.2) | 6.6 (2.4) | ns | - | - |
| PSEL (ng/mL) | 39.9 (27.4) | 41.3 (31) | 45.2 (31.5) | **<0.01** | 0.005 | SZ<HC |
| VCAM-1 (ng/mL) | 467 (143) | 480 (156) | 497 (146) | **<0.001** | 0.009 | SZ<HC |
| *IL-18 system* | | | | | | |
| IL-18 (pg/mL) | 795 (1238) | 694 (1016) | 691 (1031) | ns | - | - |
| IL-18BP (ng/mL) | 6.2 (2.6) | 5.8 (2.2) | 5.4 (2.2) | **<0.001** | 0.02 | HC<BD, SZ |
| IL-18RAP (pg/mL) | 45.5 (12.3) | 45.4 (12.7) | 46.8 (13.7) | ns | - | - |
| IL-18R1 (ng/mL) | 0.81 (0.4) | 0.8 (0.4) | 0.7 (0.4) | **0.03** | 0.003 | HC<SZ |
| *Defensins* | | | | | | |
| HNP1-3 (ng/mL) | 7 (3.2) | 6.9 (2.6) | 7.3 (4) | **<0.01** | 0.007 | BD, SZ<HC |
| BD-1 (ng/mL) | 11.3 (4.5) | 12.7 (5.4) | 11.9 (4.5) | **<0.001** | 0.01 | SZ<BD, HC; HC<BD |
| BD-2 (ng/mL) | 254 (211) | 229(207) | 199 (182) | **<0.001** | 0.01 | HC<SZ, BD |

[a]Median (interquartile range, IQR)

[b]Kruskal-Wallis test and Dunn's pairwise comparison (Bonferroni corrected)

[c]eta-squared based on H statistic: 0.01 -< 0.06 (small), 0.06 -< 0.14 (moderate), >=0.14 (large)

Abbreviations: Schizophrenia (SZ), Bipolar disorder (BD), Healthy controls (HC)

**Supplementary Table 4.** Cognitive domain scores between SZ, BD and HC

| **Cognitive domains**[a] | **SZ (N = 343)** | **BD (N = 289)** | **HC (N = 770)** | ***p*-value**[b] | **Effect size**[c] | **Pairwise comparisons**[b] |
|---|---|---|---|---|---|---|
| Fine motor speed | -0.42 (1.18) | -0.14 (1.01) | 0.31 (0.56) | **<0.001** | 0.43 | SZ<HC,BD;BD<HC |
| Psychomotor processing speed | -0.57 (0.94) | -0.02 (0.95) | 0.41 (0.83) | **<0.001** | 0.49 | SZ<HC,BD;BD<HC |
| Mental processing speed | -0.42 (1.00) | -0.05 (0.95) | 0.26 (0.73) | **<0.001** | 0.41 | SZ<HC,BD;BD<HC |
| Attention | -0.31 (0.87) | -0.16 (0.89) | 0.12 (0.94) | **<0.001** | 0.24 | SZ,BD<HC |
| Verbal learning | -0.41 (1.04) | 0.14 (0.99) | 0.27 (0.84) | **<0.001** | 0.39 | SZ<BD,HC |
| Verbal memory | -0.44 (1.09) | 0.12 (0.92) | 0.22 (0.85) | **<0.001** | 0.36 | SZ<HC,BD;BD<HC |
| Semantic fluency | -0.63 (0.86) | 0.06 (0.98) | 0.37 (0.86) | **<0.001** | 0.54 | SZ<HC,BD;BD<HC |
| Working memory | -0.42 (0.92) | -0.13 (0.90) | 0.33 (0.96) | **<0.001** | 0.41 | SZ<HC,BD;BD<HC |
| Cognitive control | -0.39 (1.09) | -0.09 (1.01) | 0.26 (0.65) | **<0.001** | 0.35 | SZ<HC,BD;BD<HC |

[a]Mean (standard deviation, SD), Z-scores

[b]Robust one-way ANOVA and lincon post-hoc for pairwise comparison (multiple comparison corrected; WRS2 R-package)[1]

[c]Explanatory measure of effect size, $\xi$ (0.1, 0.3, 0.5, small/medium/large; WRS2 R-package)

Abbreviations: Schizophrenia (SZ), Bipolar disorder (BD), Healthy controls (HC)

[1] Patrick Mair and Rand Wilcox, ‘Robust Statistical Methods in R Using the WRS2 Package’, *Behavior Research Methods*, 52.2 (2020), 464–88 (p. 2) <https://doi.org/10.3758/s13428-019-01246-w>.
